# Widespread human exposure to ledanteviruses in Uganda – a population study

**DOI:** 10.1101/2023.09.29.23296156

**Authors:** James G. Shepherd, Shirin Ashraf, Jesus F. Salazar-Gonzalez, Maria G. Salazar, Robert Downing, Henry Bukenya, Hanna Jerome, Joseph T. Mpanga, Chris Davis, Lily Tong, Vattipally B. Sreenu, Linda A. Atiku, Nicola Logan, Ezekiel Kajik, Yafesi Mukobi, Cyrus Mungujakisa, Michael V. Olowo, Emmanuel Tibo, Fred Wunna, Hollie Jackson Ireland, Andrew E. Blunsum, Iyanuoluwani Owolabi, Ana da Silva Filipe, Josephine Bwogi, Brian J. Willett, Julius J. Lutwama, Daniel G. Streicker, Pontiano Kaleebu, Emma C. Thomson

**Affiliations:** MRC-University of Glasgow Centre for Virus Research, Glasgow, UK; MRC/UVRI & LSHTM Uganda Research Unit, Entebbe, Uganda; Uganda Virus Research Institute, Entebbe Uganda; London School of Hygiene and Tropical Medicine, London, UK

## Abstract

Le Dantec virus (LDV), type species of the genus *Ledantevirus* within the *Rhabdoviridae* has been associated with human disease but has gone undetected since the 1970s. We describe the detection of LDV in a human case of undifferentiated fever in Uganda by metagenomic sequencing and demonstrate a serological response using ELISA and pseudotype neutralisation. By screening a cohort of 997 individuals sampled in 2016, we show frequent exposure to ledanteviruses with 76% of individuals seropositive in Western Uganda, but lower seroprevalence in other areas. Serological cross-reactivity as measured by pseudotype-based neutralisation was confined to *Ledantevirus,* indicating population seropositivity may represent either exposure to LDV or related ledanteviruses. We also describe the discovery of a closely related ledantevirus, Odro virus, in the synanthropic rodent *Mastomys erythroleucus*. *Ledantevirus* infection is common in Uganda but is geographically heterogenous. Further surveys of patients presenting with acute fever are required to determine the contribution of these emerging viruses to febrile illness in Uganda.

## Introduction

The epidemiology of acute febrile illness in sub-Saharan Africa is poorly characterised (1). Despite the presence of a variety of endemic human pathogens and zoonoses, the diagnostic infrastructure is limited in comparison to the extensive culture-based and molecular assays available to patients in high income countries (2). Accordingly, even in severe cases the overwhelming majority of non-malarial febrile illness in Africa is managed in line with syndromic guidelines in the absence of a microbiological diagnosis (3). Paradoxically, the existing diversity of human pathogens in sub-Saharan Africa is likely to expand further as populations encroach into novel ecosystems (4).

Although rapid diagnostic tests are widely available for malaria, diagnostics for the viral pathogens that cause febrile illness are largely unavailable outside tertiary hospitals or national referral centres. Viruses contribute heavily to common fever-related presentations in Africa (5), but many research studies perform limited viral diagnostics or omit them entirely (6). As such there is an incomplete understanding of the viruses causing febrile illness in Africa, limiting the potential for early epidemic and pandemic response. To address this, researchers have employed unbiased metagenomic Next Generation Sequencing (mNGS) for pathogen diagnosis in cohorts of patients with acute febrile illness. This approach involves amplification of all nucleic acids within a sample in a “pathogen agnostic” manner, and has discovered several novel viruses associated with human disease (7–9).

Here we describe detection of Le Dantec virus (LDV), a rhabdovirus first isolated in association with human febrile illness in Senegal in 1965, in the blood of a Ugandan patient with undifferentiated acute febrile illness in 2012 by mNGS. We demonstrate a serological response against the viral glycoprotein in patient serum by ELISA, immunocytochemistry and pseudotype neutralisation. By screening stored serum samples from a nationally representative cohort by ELISA, we show evidence of significant but geographically heterogenous exposure to LDV across Uganda and investigate the ecological factors associated with seropositivity. Testing ELISA reactive sera against a panel of related pseudotype viruses reveals that in addition to LDV, other closely related viruses are likely to be causing human infection via zoonotic transmission. In support of this possibility, we report the detection of a novel *Ledantevirus*, closely related to LDV, in a rodent host derived from peri-domestic ecological sampling in Northern Uganda.

## Results

### Patient characteristics

The findings of the Acute Febrile Illness (AFI) study including the detection of LDV have been reported previously (10). Here we focus on the clinical presentation of the LDV patient. The patient was a male child (aged under 10 years) residing in Kasese district, Western Uganda, who was recruited to the AFI study, an observational study investigating the epidemiology of acute febrile illness in Uganda. He presented to his local health centre in May 2012 with a 4-day history of fever, chills, headache, arthralgia, abdominal pain, and vomiting. He had a fever of 38.5°C, but no other abnormal findings on physical examination. A presumptive diagnosis of typhoid was made by the treating physician based on clinical features and he was managed as an outpatient with oral ciprofloxacin. He had fully recovered at a follow-up appointment four weeks later and at further follow up in 2018 he remained well. After enrolment in the AFI study, blood samples were tested against multiple pathogens including leptospirosis, brucellosis, malaria, dengue, chikungunya, yellow fever, o’nyong nyong, typhoid and rickettsiosis, all of which were negative (Table S1).

### Detection of LDV by metagenomic sequencing of acute patient samples

As initial diagnostic investigations were negative, a serum sample from the acute phase of the patient’s illness was subjected to mNGS. Extracted RNA from serum was sequenced on an Illumina MiSeq instrument, resulting in 1677574 paired-end sequencing reads. *De novo* assembly yielded a 11,444 nucleotide contig that aligned to the genome of LDV on blastn query of the NCBI nucleotide database (isolate DakHD763, KM205006, 11450 base pairs). This contig was used as a reference sequence for alignment of raw sequencing reads, resulting in alignment of 8958 reads with 99.7% genome coverage at a minimum depth of 10 reads. Pairwise alignment of the final genome showed greater than 90% nucleotide similarity to the original LDV isolate DakHD763 in all genomic regions (Table S2). Phylogenetic analysis confirmed the placement of the Ugandan LDV isolate within the genus *Ledantevirus* (Figure 1).

**Figure 1.**
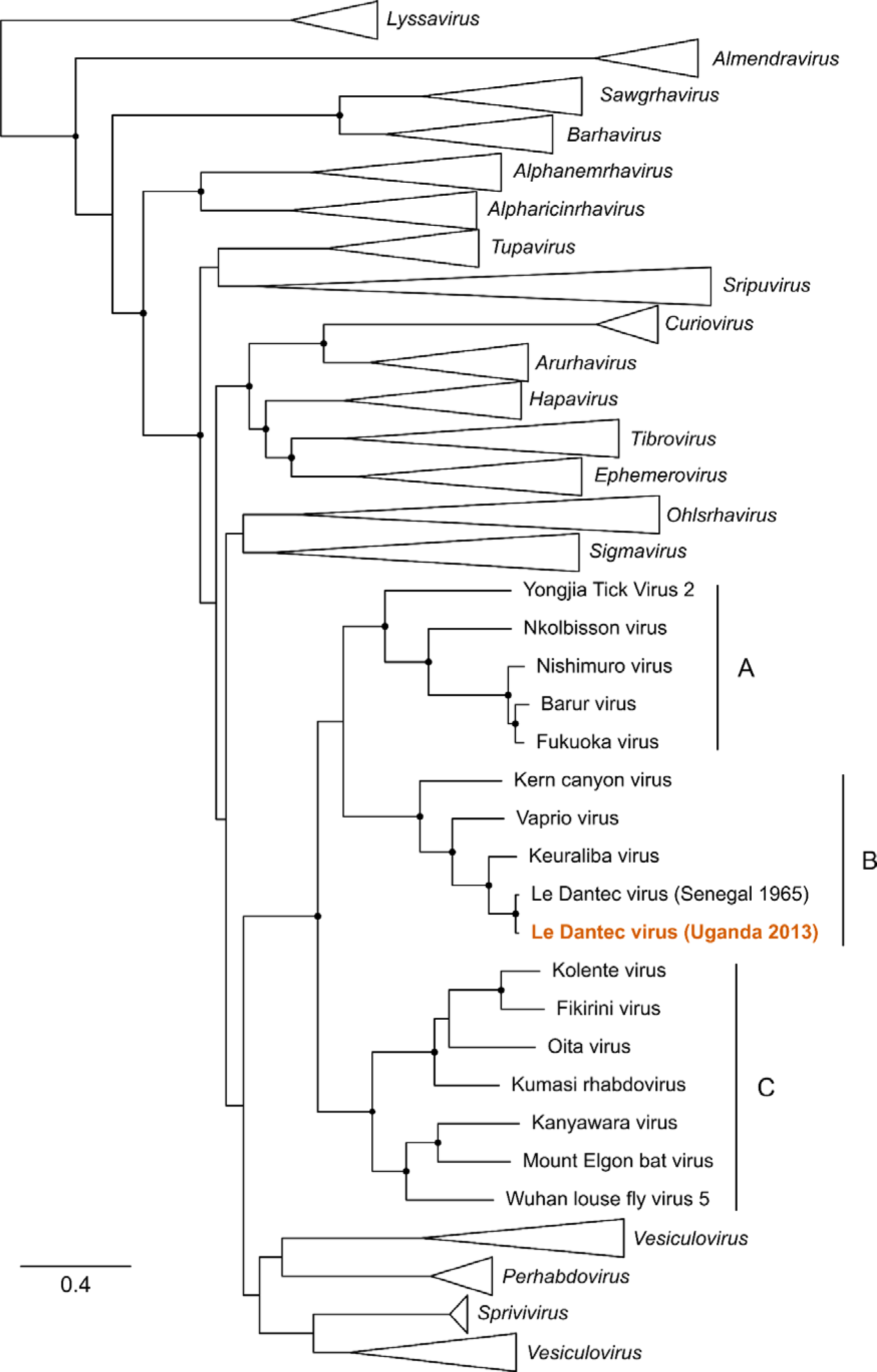
Phylogenetic relationships of Le Dantec Virus identified in Uganda. Maximum likelihood phylogeny of the sub-family *Alpharhabdovirinae* based on full-length L protein amino acid sequences. The genus *Ledantevirus* is expanded with phylogroups A, B and C indicated to the right. Other genera are collapsed. The sequence derived from the febrile patient described in this study is indicated in orange text. The scale bar represents substitutions per site. Nodes with bootstrap support >70 are indicated with black circles.

### Confirmation of LDV infection by PCR and screening of acute fever samples

To confirm mNGS discovery of LDV, a freshly prepared RNA extract from patient serum was assayed by RT-PCR with primers designed against conserved nucleotide positions in the original DakHD763 isolate and the sequence derived from Ugandan patient. The PCR assay yielded a 5.4 Kb fragment. A separate RT-PCR assay targeting a 2.4 Kb fragment in the glycoprotein gene was also positive. Sanger sequencing of the 5.4 Kb amplicon revealed all but one of the 5,406 nucleotides to be identical to the metagenomic sequence, validating the metagenomic sequence and confirming the presence of LDV RNA in the serum of the Ugandan patient.

The RNA was also tested with a separate LDV-specific real-time RT-PCR assay. Using serial 10-fold dilutions of the DakHD763 isolate RNA as positive control we detected the 145-bp LDV sequence with a linear dynamic range of over four orders of magnitude. A fresh RNA extract from the source patient also showed a detectable signal; three µl of plasma-equivalent RNA yielded a Ct value of 30, consistent with a substantial viremia. Plasma samples (n=62) from a measles/rubella cohort and plasma pools (10 per pool) of unexplained acute febrile infections from Sudan tested all negative for LDV RNA in the real-time RT-PCR assay.

### Demonstration of a serological response to infection with LDV in the index patient

Using an in-house LDV glycoprotein (LDV-G) ELISA we demonstrated reactivity to LDV-G by both purified IgG and serum from blood samples collected from the patient at a follow-up visit in 2018 (Figure 2). Reactivity was also detected in a close relative who had experienced a contemporaneous illness but had not presented to hospital. A further 10 unrelated serum samples from Uganda and 12 from healthy UK controls were tested, with evidence of an antibody response to LDV-G detectable in both the index case and their close contact as well as two individuals from Uganda and one sample from the UK (UK10). On further investigation it emerged that UK10 had previously resided in Africa.

**Figure 2.**
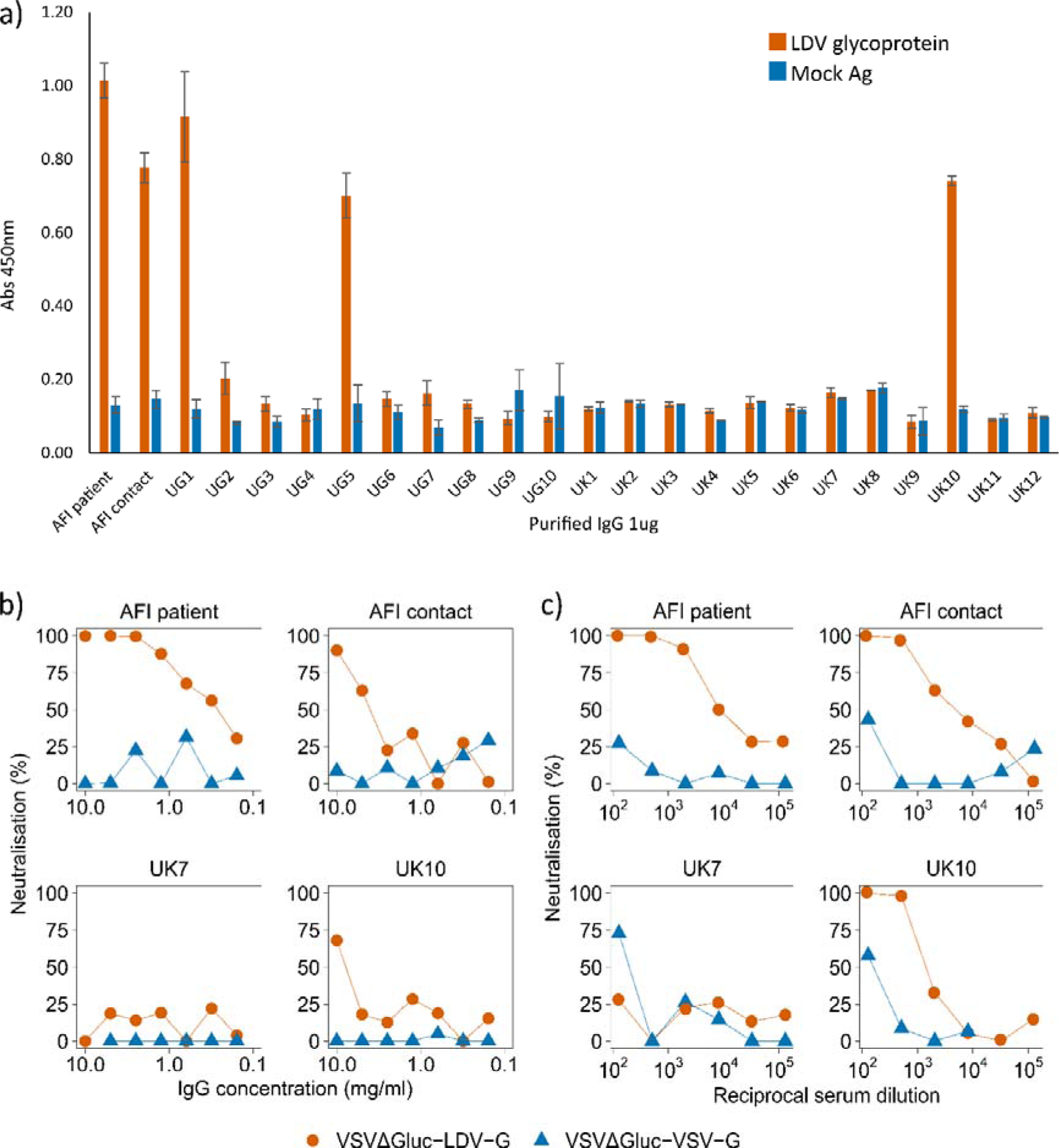
Serological detection of LDV in patient serum by ELISA and pseudotype neutralisation. a) Raw OD450 values for purified IgG from the index patient, a close contact and Ugandan and UK controls tested by LDV-G ELISA. Bars represent the mean OD450 based on six technical replicates. Error bars represent the standard error of the mean. b) Neutralisation of VSVΔG*luc*-LDV-G and VSVΔG*luc*-VSV-G by purified IgG from human serum. Serial IgG dilutions were mixed with pseudotype preparations and added to HEK293T cells. Luciferase activity was measured at 48 hours post infection. Datapoints represent the mean neutralisation derived from three technical replicates. c) Neutralisation of VSVΔG*luc*-LDV-G and VSVΔG*luc*-VSV-G by human serum. Serial serum dilutions were mixed with pseudotype preparations and added to HEK293T cells. Luciferase activity was measured at 48 hours post infection. A representative experiment is shown. Datapoints represent the percentage neutralisation derived from the mean of three technical replicates. Percentage neutralisation represents luciferase readings relative to no-serum control wells.

A vesicular stomatitis virus (VSV) pseudotype virus with the glycoprotein gene replaced with the gene for firefly luciferase (VSVΔG*luc*) was created to express LDV-G as an outer membrane protein (VSVΔG*luc*-LDV-G). Both serum and purified IgG from the febrile patient specifically inhibited VSVΔG*luc*-LDV-G, without a similar effect on pseudotype viruses displaying the VSV glycoprotein (VSVΔG*luc*-VSV-G, Figure 2b). Consistent with the LDV-G ELISA, serum and purified IgG from the close family contact and UK10 inhibited VSVΔG*luc*-LDV-G, although to a lesser degree than the patient.

To visualise the interactions between LDV-G and patient IgG, LDV-G engineered with a 6-histidine C-terminal tag (LDV-Ghis) was expressed in BHK-21 cells. Cells were incubated with a mixture of a rabbit anti-histidine primary antibody and patient sera. Co-localisation of the rabbit anti-histidine and IgG from patient samples was demonstrated for the patient and other samples found to be positive by ELISA, further demonstrating a serological response directed against LDV-G (Figure 3).

**Figure 3.**
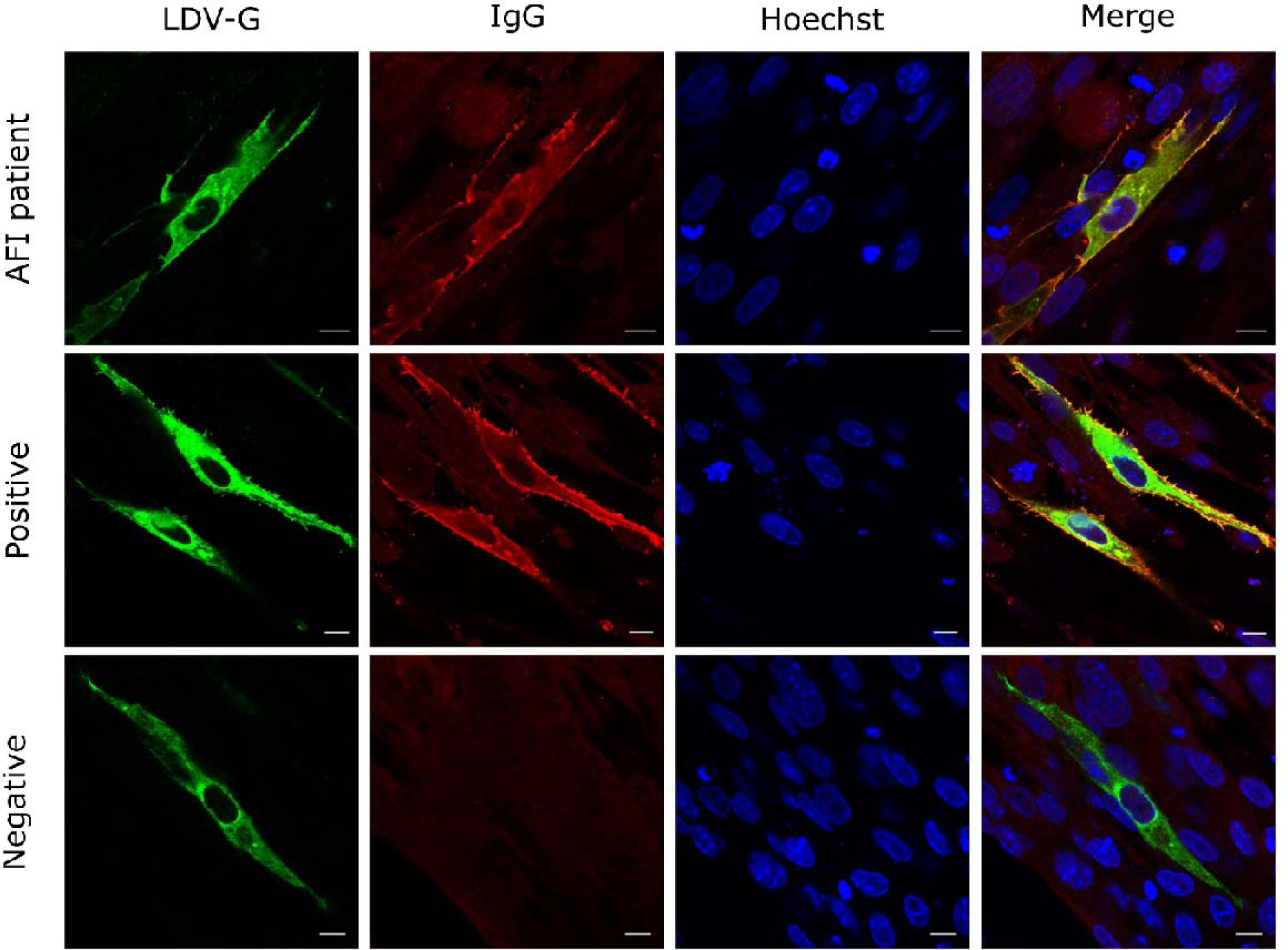
Co-localisation of LDV-G and human IgG by immunocytochemistry. BHK-21 cells were transfected with the mammalian expression plasmid VR1012-LDV-Ghis then fixed and permeabilised. Primary antibodies were a rabbit anti-his and human serum samples diluted 1:200. Secondary antibodies were an Alexa Fluror 488 conjugated anti-rabbit antibody (green) and anti-human 594 (red). Each sample was tested in duplicate. A representative experiment including the Ugandan patient serum and positive and negative samples as tested by LDV-G ELISA is shown. Three biological replicates were performed for each sample, with the exception of sera from the index patient where a single replicate was performed owing to limited sample availability.

### High population exposure to LDV in Uganda

To investigate exposure to LDV in the Ugandan population, we screened a cohort of stored serum samples collected in 2016 as part of the Ugandan national HIV antenatal seroprevalence study (ANC-2016) by LDV-G ELISA. The cohort comprised a random selection of sera from 997 women of childbearing age (median age 25, IQR 21-29, range 15-48) from 24 locations across Uganda (Figure 4).

**Figure 4.**
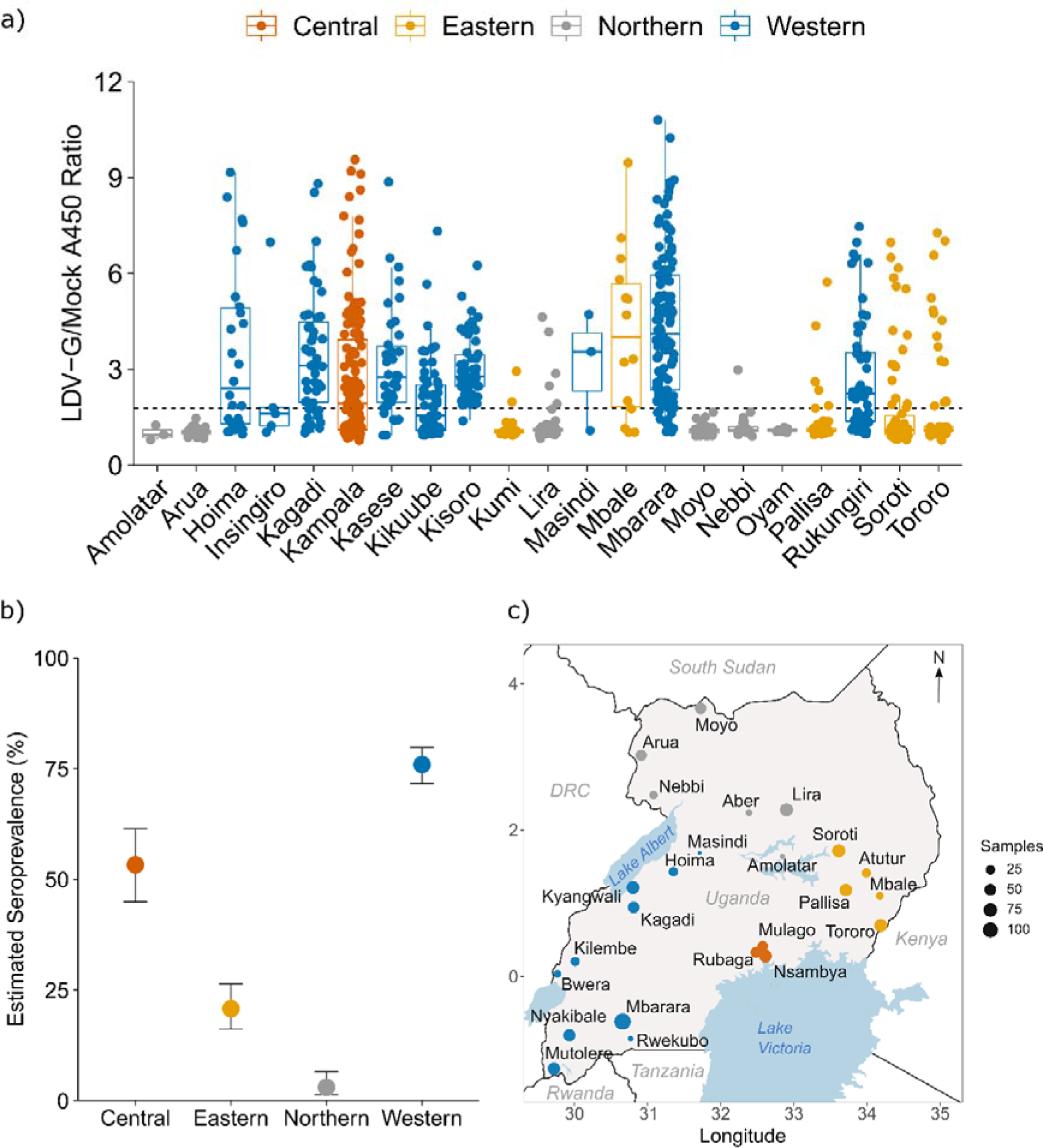
Seroprevalence of ledanteviruses in Uganda by LDV-G ELISA. (a) Raw A450 index values by district of Uganda derived from in-house LDV-G capture ELISA. Colours represent the regions of Uganda. A450 ratio values were derived by dividing the absorbance value of the test well by that of the paired mock well for each sample. Data points represent the mean value for a single individual derived from three independent experiments. Centre bars represent the median, box edges the IQR, and vertical lines the range of values within 1.5 times the interquartile range. The dashed line represents the positive cut-off derived from the mean + 3 × the standard deviation of samples derived from the West Nile subregion. (b) Estimated seroprevalence of LDV across the major regions of Uganda based on in-house LDV-G ELISA. Seroprevalence is displayed as the proportion of positive cases +/-95% confidence intervals based on the following sample sizes: Central n=137, Eastern n=240, Northern n=196, Western n=424. (c) map of Uganda and surrounding countries and major waterbodies showing locations of study sites from the ANC-2016 study. Coloured circles represent locations of study sites with the size of each circle corresponding to the number of participants from each site tested by the LDV-G ELISA.

There was heterogeneity in reactivity of serum samples by LDV-G ELISA between the four administrative regions of Uganda (Figure 4). Notably, samples from Northern sites demonstrated low reactivity, in particular the sites in the West-Nile sub-region; Arua, Moyo and Nebbi. As these sites clustered spatially and shared a significantly lower reactivity when compared with other regions, they were considered to represent a low-prevalence population and were used to estimate a positive cut-off value for the assay (mean A450 ratio + 3 standard deviations). Applying this cut-off to all samples we estimated seroprevalence for the different regions of Uganda, with exposure highest in the Western region (75.9%, 95% CI: 71.7-79.8), followed by the Central region (53.3%, 95% CI: 44.9 - 61.4) and the Eastern region (20.8%, 95% CI: 16.2 - 26.4). Exposure was markedly lower in the Northern region (3.1%, 95% CI: 1.4 - 6.5, Figure 4b). The only patient-level metadata recorded for the ANC-2016 dataset was age, which was significantly associated with seropositivity on univariate analysis (Mann-Whitney U, p=0.016, Figure S1).

### Cross-reactivity of LDV-G reactive sera between phylogroup B ledanteviruses

LDV antisera cross-reacts with Keuraliba virus (KEUV) in complement fixation assays (11, 12). We therefore investigated whether reactivity to LDV-G by ELISA could be explained by serological cross-specificity to related viruses rather than exposure to LDV. We generated VSVΔG*luc* pseudotypes expressing the glycoproteins of three representative phylogroup B ledanteviruses; KEUV, Vaprio virus (VAPV) and Kern Canyon Virus (KCV), with pseudotypes expressing VSV glycoprotein as a negative control. First we determined IC50 values for sera from the index patient against each pseudotype. Consistent with prior infection serum titres were highest against VSVΔG*luc*-LDV-G (IC50=6864), followed by VSVΔG*luc*-KEUV-G (IC50=2981), VSVΔG*luc*-KCV-G (IC50=983) and VSVΔG*luc*-VAPV-G (IC50=141). There was minimal neutralising activity against VSVΔG*luc*-VSV-G (IC50=71).

Next, we randomly selected sera from individuals in the ANC-2016 cohort that were positive by LDV-ELISA and determined serum IC50 values against the pseudotype viruses. There was significant cross-neutralisation of pseudotypes bearing LDV-G and KEUV-G by ELISA positive sera (Figure 5a, 5b). Notably, IC50 titres against KEUV-G were in general higher than those for LDV-G (median LDV-G IC50 2121, median KEUV-G IC50 3166.4, median IC50 fold change for KEUV-G relative to LDV-G 0.52, IQR: 0.24-1.14). There was cross-neutralisation to a lesser extent of VAPV-G (median IC50 to VAPV-G 457.2, median fold change relative to LDV-G = 6.09, IQR: 2.26 – 9.27) and KCV-G (median IC50 against KCV-G 252, median fold change relative to LDV-G = 9.32, IQR = 4.73-18.1). As expected, pseudotypes bearing VSV-G were neutralised to the lowest extent (median IC50 106.4, IQR: 74.7-178.1, median fold change relative to LDV-G = 17.5, IQR: 8.21-32.5). To determine if there was an obvious spatial pattern differentiating the LDV-G ELISA positive sera based on their IC50 against LDV-G or KEUV-G, the LDV/KEUV fold change in neutralisation was expressed as log2 fold change (Figure 5c). There was no obvious geographical pattern consistent with spatial restriction of sera with higher IC50 to either LDV or KEUV.

**Figure 5.**
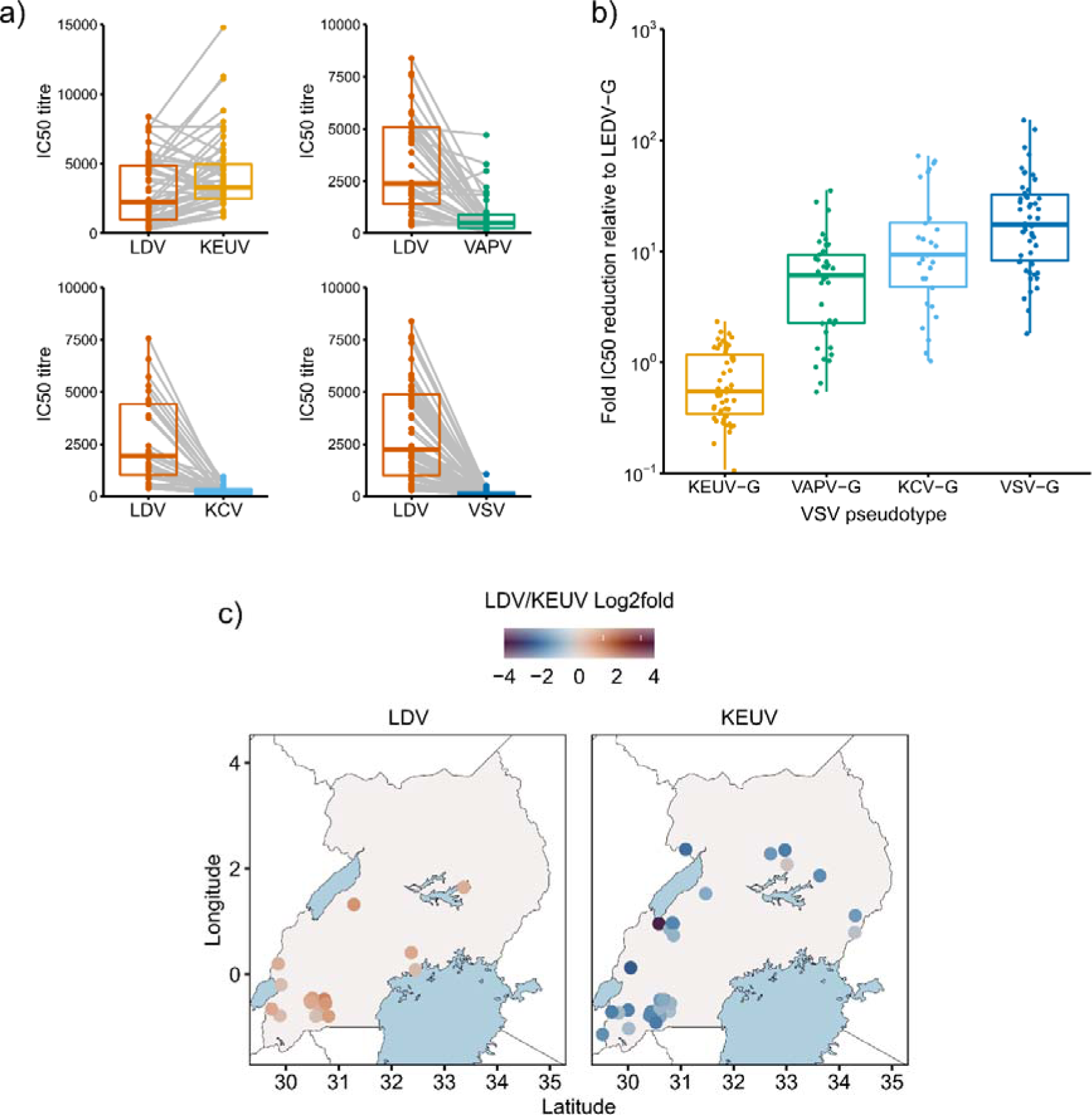
LDV-G ELISA positive serum exhibits serological cross-reactivity within *Ledantevirus*. a) IC50 titres by viral pseudotype for ANC-2016 serum samples positive by LDV-G ELISA. Between group differences were tested by paired Wilcoxon test. a; n = 60, median LDV IC50 2121, median KEUV IC50 3166.4, p < 0.001. b; n = 46, p <0.001. c; n = 37, median LDV IC50 1863, median KCV IC50 252, p <0.001. d; n = 60, p <0.001. Centre bars represent the median, box edges the IQR, and vertical lines the range of values within 1.5 times the interquartile range. Data points represent the IC50 titre of each patient sample as determined by 4-parameter logistic regression curves derived from 2 biological replicates. Grey lines link the titres for an individual against different pseudotypes. B) Fold reduction in IC_50_ titre of sera against each VSVΔG*luc* pseudotype relative to VSV(LDV-G). VSV(KEUV-G) (n = 52, median = 0.54, IQR 0.33-1.14), VSV(VAPV-G) (n = 38, median = 6.09, IQR = 2.26–9.27), VSV(KCV-G) (n = 29, median = 9.32, IQR = 4.73-18.1) and VSV(VSV-G) (n = 52, median = 17.5, IQR = 8.21-32.5). Significance was determined by Kruskall-Wallis rank-sum test (H = 111.31, df = 3, p < 0.001). c) Relative distribution of sera neutralising major *Ledantevirus* species in Uganda. Colour intensity represents log2 fold change in neutralising titre (LDV/KEUV IC50) for ELISA positive samples. Panel A; Samples with LDV titre greater than KEUV (n = 18). Panel B; Samples with KEUV titre greater than LDV (n=34). Data points represent individual patients. Exact locations are jittered to avoid overlaps.

### Environmental correlates of LDV-G seropositivity in Uganda

As there was evidence of significant regional variation in seropositivity by LDV-G ELISA within Uganda, the influence of environmental variation at the ANC-2016 study sites was investigated by general linear mixed modelling (GLMM). Climatic (relating to rainfall and temperature), geographical (elevation and tree cover) and livestock density variables were determined for each study site. Multicollinear variables were identified and excluded and a binomial GLMM constructed with the effect of recruitment site controlled by its inclusion as a random effect. The age of individual participants was included as an explanatory variable. Model selection based on AICc showed patient age (OR 1.04 p = 0.004), forest cover (OR 1.89 p = 0.016) and the climatic variable isothermality, which describes daily and seasonal temperature fluctuations, (OR 4.81 p <0.001) were positively associated with seroprevalence (Table 1). Notably, no effect of livestock density was observed.

**Table 1.** GLMM of LDV-G seropositivity by patient age and ecological variables.

|  |  |  |  |  |  |  |
| --- | --- | --- | --- | --- | --- | --- |
|  | Estimate | Std. err | Z value | Pr(>\|z\|) | OR | 95% conf int. |
| (Intercept) | -1.59483 | 0.45667 | -3.492 | <0.001 | 0.21 | 0.08 – 0.50 |
| Age | 0.04232 | 0.01472 | 2.875 | 0.004 | 1.04 | 1.01 – 1.07 |
| Forest cover | 0.63832 | 0.26530 | 2.406 | 0.016 | 1.89 | 1.10 – 3.33 |
| Isothermality | 1.57128 | 0.31730 | 4.952 | <0.001 | 4.81 | 2.63 – 9.93 |
| Marginal $R^2$: 0.466, Conditional $R^2$: 0.596 | | | | | | |

### Environmental evidence of novel ledanteviruses

Whilst LDV has only been detected in humans, KEUV was originally isolated from various rodent species in Senegal (11). To investigate ecological reservoirs of viral pathogens in Uganda, we performed live rodent trapping in Arua district and used mNGS to investigate the blood virome of captured rodents. Over 2233 trap nights we captured 205 rodents (Table S3). We detected genomic evidence of a novel rhabdovirus, which we have named Odro virus, in blood from a multimammate mouse, *Mastomys erythroleucus,* trapped in Arua district in 2019. Host species was confirmed by sequence determination of the mitochondrial cytochrome B gene (Figure S2). Sequencing yielded 1383706 reads of which 584 mapped to viral species within *Ledantevirus* by blastx. Target enrichment sequencing of the NGS library yielded a further 1296041 reads, of which 11301 mapped to *Ledantevirus*. *De novo* assembly of the combined reads from metagenomic and target enriched sequencing yielded 13 contigs corresponding to fragments of the nucleoprotein, phosphoprotein, glycoprotein and RDRP of a rhabdovirus with high similarity to members of *Ledantevirus* (Table S4). Further iterative *de novo* assembly using these contigs as scaffolds yielded 88% coverage of the putative nucleoprotein, 75% of the phosphoprotein, 72% of the glycoprotein, and 89% of the RDRP with reference to the closest related sequence on the NCBI database. Phylogenetic analysis placed these fragments into phylogroup B of *Ledantevirus*, alongside LDV and KEUV (Figure 6). Pairwise amino acid distances in the G and L proteins suggest Odro virus is a novel species within *Ledantevirus* (Table S5). In addition to coding sequences, there was adequate sequencing coverage of gene junction sequences for the N-P, P-M and U-L intergenic regions, demonstrating the conserved rhabdovirus intergenic transcription termination-polyadenlyation (3’-ACUUUUUUU-5’), and transcription initiation sequences (3’-UUGUnnUAG-5’) (13). Consistent with other phylogroup B ledanteviruses, the polyadenylation signal at the P-M junction incorporates the termination codon of the upstream phosphoprotein ORF (Supplementary Figure S3).

**Figure 6.**
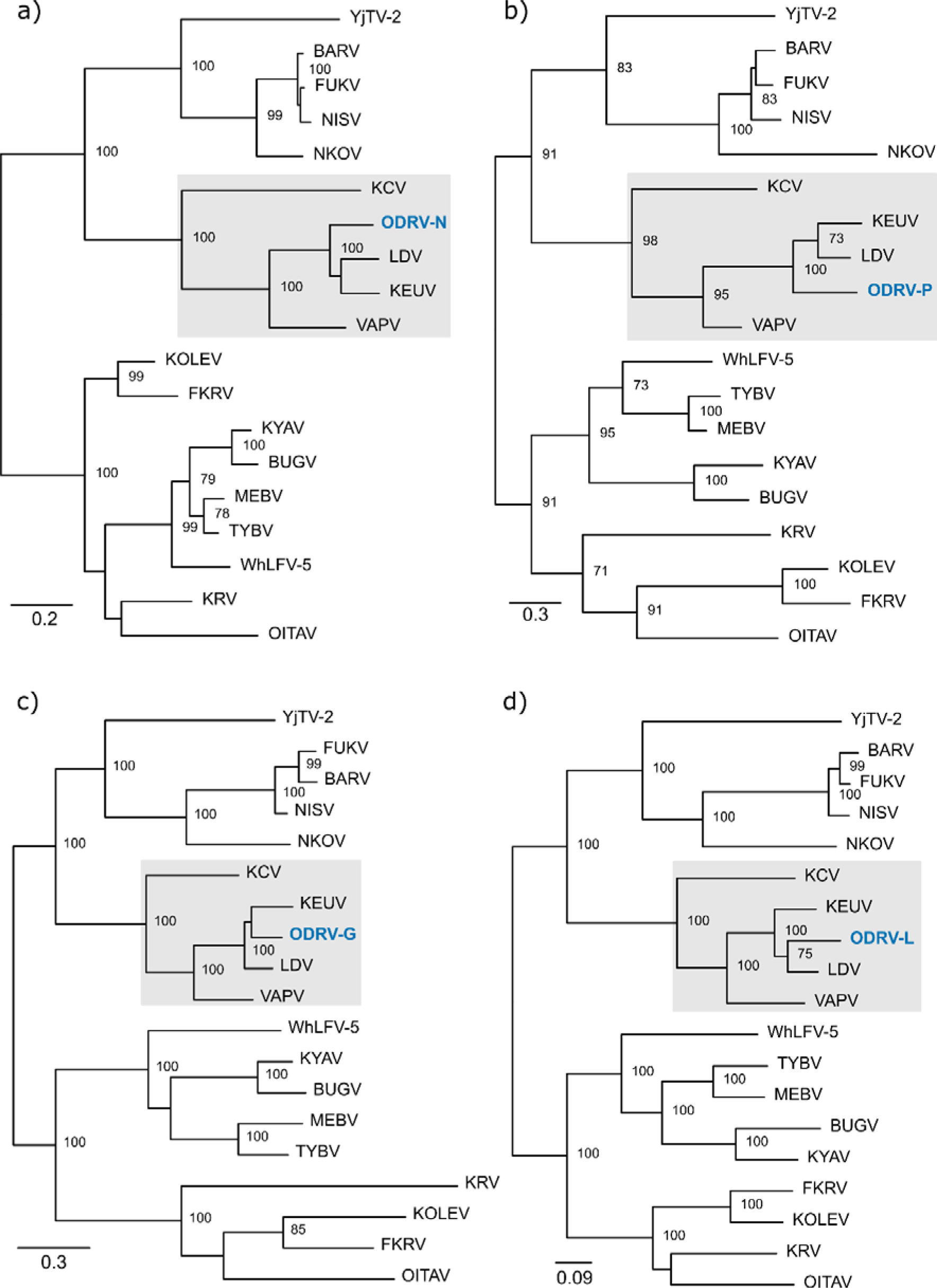
Phylogenetic placement of Odro virus genomic fragments within *Ledantevirus*. (a) a continuous 373 amino acid fragment corresponding to the Odro virus nucleoprotein. (b) three continuous fragments of 87aa, 72aa and 73aa corresponding to the Odro virus phosphoprotein. (c) a continuous 411 amino acid fragment corresponding to the Odro virus glycoprotein. (d) five fragments of 65aa, 120aa, 529aa, 814aa and 168aa corresponding to the Odro virus L protein. For each gene alignments were created based on complete coding sequences of all members of *Ledantevirus* and gene fragments of Odro virus and maximum likelihood phylogenies generated. Node labels represent bootstrap support values for nodes with support >70 based on 1000 replicates. The grey highlighted areas represent the phylogroup B viruses. ODRV; Odro virus. LDV; Le Dantec virus, KEUV; Keuraliba virus, VAPV; Vaprio virus, KCV; Kern Canyon virus, YjTV-2; Yongjia tick virus 2, BARV; Barur virus, FUKU; Fukuoka virus, NISV; Nishimuro virus, NKOV; Nkolbisson virus, WhLFV-5; Wuhan louse fly virus 5, TYBV; Taiyi bat virus, MEBV; Mount Elgon bat virus, BUGV; Bughendera virus, KYAV; Kanyawara virus, FKRV; Fikirini rhabdovirus, KOLEV; Kolente virus, KRV; Kumasi rhabdovirus, OITAV; Oita virus.

## Discussion

We describe the second confirmed case of human infection with LDV and demonstrate evidence of high population exposure to LDV or closely related ledanteviruses in Western Uganda. In addition, we report genomic evidence of Odro virus, a novel ledantevirus, in blood from a *Mastomys erythroleucus*, suggesting a rodent reservoir of African ledanteviruses. *Ledantevirus* is an expanding genus within the family *Rhabdoviridae*, with 18 members designated in the latest ICTV taxonomy update (14). The genus is divided into three phylogroups based on molecular similarity. In contrast to the related genera *Vesiculovirus* and *Sigmavirus*, whose members are predominantly associated with invertebrates, most ledanteviruses have been isolated from vertebrate hosts, or from ectoparasites such as bat flies (15, 16). LDV was originally isolated from a 10-year-old female who presented to hospital in Senegal in 1965 with fever (17). In addition to LDV, two other members of *Ledantevirus* have a reported association with human infection. Serological evidence of exposure to Kumasi virus was demonstrated in individuals residing in the area in which it was isolated from a bat in Ghana (18). Nkolbisson virus, originally isolated in 1964 from mosquitoes in Cameroon, was subsequently reported to have been isolated from human samples in the Central African Republic, although there are few details available of its association with human disease (19, 20).

Metagenomic sequencing has been increasingly applied in cases of unidentified acute febrile illness in Africa, leading to detection of several novel viruses including rhabdoviruses (9, 21). Determining the clinical significance of viral species detected by sequencing alone can however be challenging (22). Demonstration of seroconversion or detection of viral antigens in diseased tissue by histology can support the role of a novel agent as a pathogen (23, 24), but the clinical samples required for such analyses are often unavailable and can be challenging to access in low-resource settings. Tissue sampling for histological examination is particularly difficult to obtain in non-fatal cases. Thus, it is often unclear if sporadically detected agents such as LDV represent pathogens or are incidental to an alternative disease process. The recent detection of two novel orthobunyaviruses in Uganda, both clearly associated with febrile illness but for which serological or other pathological parameters have not yet been reported, are illustrative of this challenge (7, 8). Systematic screening for novel or emerging pathogens by mNGS at sentinel clinical sites or through national surveillance systems may allow accurate estimation of their contribution to local infectious disease epidemiology, and inform the need for development of diagnostic platforms for use beyond research settings.

LDV has been exclusively detected in association with febrile illness and can be classed as a probable pathogen. To support this, we have shown a specific serological response by the patient directed against LDV-G, although owing to unavailability of samples from the acute phase of illness we were unable to demonstrate seroconversion. The patient tested negative against a panel of pathogens commonly causing febrile illness in this setting. Additionally, we report widespread population exposure to either LDV itself or other members of *Ledantevirus* in Uganda, particularly in the Western Region. The high seroprevalence in this area suggests ledanteviruses may predominantly cause subclinical or mild infection and that more severe cases such as that described here go unreported or undiagnosed. Notably the patient in this case made a full recovery. Such viruses are often able to spread undetected by surveillance systems in low and middle-income countries as demonstrated by the emergence and expansion of Zika virus (25). Even if resultant disease is in most cases non-severe, pathogens can exert significant pressure on health systems and cause substantial economic effects if transmission within human populations increases. Patients in Uganda are able to purchase antibiotics without a prescription and individuals may self-medicate at the onset of symptoms before presenting to healthcare if they fail to improve (26). Circulation of viral pathogens causing even mild, self-limiting febrile illness thus serves to drive inappropriate antimicrobial use and antimicrobial resistance. As we detected LDV in a sample originally collected in 2012, and our serological data is based on samples from 2016, further prospective studies are required to determine the current contribution of ledanteviruses to AFI in Uganda.

Most of the LDV-G ELISA positive sera in the ANC-2016 cohort tested against a panel of ledantevirus glycoproteins had higher IC50 titres against KEUV-G than LDV-G, suggesting that in addition to LDV, human infection by closely related ledanteviruses may be occurring in Uganda. Serological cross-reactivity between LDV and KEUV was first noted in the 1980s (11), with the close serological relationship mirrored by relatively high similarity at the amino acid level (17). A reservoir species for LDV has not yet been identified, but KEUV was initially isolated from various rodent genera in Senegal in 1968 including *Mastomys* and gerbils of genus *Taterea* and *Taterillus*. Consistent with the serological data suggesting a further member of the LDV serogroup is causing human infection in Uganda, we detected the partial sequence of a close relative of KEUV and LDV by mNGS of blood sampled from a *Mastomys erythroleucus* in Arua district. The Odro virus glycoprotein displayed 76.7% amino acid identity with KEUV and 75.6% with the LDV glycoprotein. This putative novel *Ledantevirus* is therefore likely to exhibit cross-reactive behaviour in serological assays with both LDV and KEUV and its presence in a reservoir species known for its close interaction with humans potentially explains the high seroprevalence based on LDV-G ELISA observed for some regions in the ANC-2016 cohort. Unfortunately our study is limited by our inability to determine the complete Odro virus glycoprotein sequence, meaning we were unable to test for evidence of exposure in the ANC-2016 cohort by pseudotype-based neutralisation.

Ledantevirus seroprevalence as measured by LDV-G ELISA was geographically heterogenous within Uganda, suggesting either regional variation in the presence or prevalence of infection within reservoir species or differences in the dynamics of human exposure. Notably, Odro virus, a potential driver of human LDV seroprevalence in Uganda, was discovered in Arua district where human LDV-G seroprevalence was low. One explanation is that other ledanteviruses, not present in Arua and other low seroprevalence regions, are contributing to human infection. However, if Odro virus is implicated in seropositive cases not attributable to LDV, factors other than the presence of the reservoir host are likely important in mediating the extent of human exposure. As an example, the practice of hunting of wild animals for food carries a risk of exposure to zoonotic pathogens. Regional variation in bushmeat consumption exists owing to local preferences, the availability of wildlife species, and poverty (27, 28). Rodents, particularly *Mastomys*, are often hunted by male children and adults (29, 30), highlighting a limitation of our use of an all-female adult cohort for seroprevalence.

The ecological sampling reported in this study was relatively limited in scale, and further study of wildlife in Uganda is required to accurately determine the host range of Odro virus. *Mastomys* species are important vectors of zoonoses in Africa, and are primarily found in peri-domestic or bush settings in Arua district rather than in huts or other human habitations, having been replaced as a domestic species by the expansion of *Rattus rattus* into Africa throughout the 20^th^ century (29, 31). Our analysis of ecological conditions suggests forest cover and isothermality are associated with increased human exposure to ledanteviruses in Uganda. *Mastomys* species are known to breed and carry litters throughout the year in the presence of favourable ecological conditions and the absence of drought (29, 32). It may be hypothesised that the tendency for increased bushmeat consumption in areas adjacent to forested areas, combined with stable climatic conditions that permit higher rodent densities throughout the year could act to increase population exposure to zoonotic disease in these areas. Alternatively, isothermality and forest cover may simply be correlated with the presence of a greater diversity of wildlife, and an as yet unsampled species may be driving the higher seropositivity in humans in these regions.

In summary we identified LDV as the likely causative agent in a case of acute febrile illness in Uganda and demonstrated widespread exposure to ledanteviruses in Western, Central and Eastern regions of the country with evidence of increased exposure with age and in regions with high isothermality and forest cover. A closely related novel phylogroup B *Ledantevirus* was detected in a common and geographically widespread rodent species. Identification of the infectious agents which routinely spill-over into human populations, providing opportunities for emergence and evolution within human hosts is important for pandemic preparedness. Further characterisation of the epidemiology of acute febrile illness and population level serosurveys are required to establish the precise contribution of ledanteviruses to human disease in Uganda. Our detection of a novel ledantevirus in a *Mastomys*, together with the earlier detection of KEUV in rodents, is supportive of a rodent reservoir of African ledanteviruses, which should be confirmed by further study of both rodent and non-rodent hosts in areas with evidence of human exposure.

## Methods

### Ethical approvals

Ethical approval for the AFI study, the use of stored serum samples from the ANC-2016 cohort and for wild rodent collections in Arua district was granted by the Research Ethics Committee of the Uganda Virus Research Institute (reference numbers GC/127/10/02/19, GC/127/18/09/662 and GC/127/19/06/662). Research approvals were granted by the Uganda National Council for Science and Technology (UNCST, reference numbers HS2485 and HS767).

### Clinical samples

Acute serum from the patient was collected as part of the acute febrile illness study (10). Recruitment to the study required written informed consent. A further episode of sampling for additional sera from this individual occurred in 2018. Samples from the ANC-2016 HIV surveillance study were collected in 2016 from individuals who had provided informed consent for the future use of their samples for medical research. ANC-2016 samples were supplied by Robert Downing (UVRI).

### Rodent sampling

Rodent sampling and euthanasia was conducted in line with international best practice guidelines (33). Rodent trapping was performed in three villages in Adumi subcounty; Ombaci, Oniba, and Sua, in April 2019. Rodents were trapped in Sherman traps (H.B. Sherman Trap Company) and Tomahawk traps (Tomahawk Live Trap Company) baited with peanut butter mixed with smoked fish and a piece of sweet potato. Traps were set in four locations in relation to homesteads: indoors (inside sleeping or cooking huts), the peri-domestic compound area (within five metres of the sleeping and cooking huts), the peri-domestic bush (within five metres of the homestead edge), and sylvatic (up to 300 metres into the bush from the centre of the village). Two Sherman and two Tomahawk traps were placed within the peri-domestic compound area. An additional three Sherman and three Tomahawk traps were placed in the peri-domestic bush setting. To sample the sylvatic environment one Tomahawk and one Sherman trap were placed every 20 metres for 300 metres from the centre of the village in each of the four cardinal directions (North, East, South, West).

Traps were set before dusk and opened shortly after dawn. Trapping was conducted over three nights in Ombaci and Oniba and for two nights in Sua for a total of 2233 trap nights. Seven traps were lost during the sampling period. Rodents were euthanised in the field by inhalation of halothane and cervical dislocation, followed immediately by blood sampling by heart puncture. Animals were speciated using taxonomic keys. Blood samples were centrifuged and sera heat inactivated and stored at −80 °C until further use.

### Next generation sequencing

For human and rodent serum samples RNA was extracted from 200 µl of plasma using the Agencourt RNAdvance Blood Kit (Beckman Coulter) according to the manufacturer’s instructions and eluted in nuclease-free water. RNA was reverse transcribed using Superscript III (Invitrogen) followed by dsDNA synthesis with NEBNext (New England Biolabs). DNA libraries were prepared using LTP low-input Library preparation kit (KAPA Biosystems). Resulting libraries were quantified with the Qubit 3.0 fluorometer (Invitrogen) and their size determined using a 4200 TapeStation (Agilent). Libraries were pooled in an equimolar ratio and sequenced on the Illumina MiSeq platform using 150×2 v2 cartridges.

Target enrichment of RNA derived from rodent blood was performed using custom-designed biotinylated RNA probes targeting full genomes of all known arboviruses, including *Ledantevirus* (Agilent) using manufacturer’s instructions. Briefly, dual-indexed DNA library was incubated overnight at 65°C. Captured DNA was recovered using Streptavidin T1 Dynabeads, washed with provided buffers, and eluted in water. Captured library was amplified using Illumina primers and sequenced on an Illumina NextSeq 500.

### Bioinformatic analysis

Raw sequence reads were classified using diamond version 0.8.20.82 in blastx mode (34) and visualised using Krona (35). *De novo* assembly of the human serum sample was performed with SPAdes version 3.11.1 (36). The *de novo* contigs were screened using diamond blastx and confirmed by blastn against the NCBI nr database. The *de novo* assembly was then used as a reference to align raw sequencing reads using the short read aligner tanoti (https://github.com/vbsreenu/Tanoti). For sequencing of rodent serum denovo assembly was performed using the metavic pipeline (37) and contigs mapping to *Ledantevirus* species were determined by diamond blastx. The genome organisation of *Ledantevirus* contigs was manually determined by approximating their position based on their alignment by blastx against either the LDV or KEUV reference genomes. The resulting construct was used as a scaffold for further iterative *de novo* assembly with Gapfiller version 1-11 (38). Cytochrome B sequence barcoding for speciation of *Mastomys* was performed by aligning NGS sequencing reads to the optimal *Mastomys* mitochondrial reference sequences, with the optimal reference determined by blastn query of initial consensus sequence against the NCBI nucleotide database (MZ131552). The subsequent consensus was aligned with a panel of previously published reference sequences (39).

### Phylogenetic analysis

To determine the phylogenetic placement of the LDV-Uganda L protein sequence recovered from Ugandan patient, full-length L protein amino acid sequences from representative rhabdovirus taxa within *Alpharhabdovirinae* were aligned. For phylogenetic placement of partial amino acid sequences of the putative N, P, G and L genes of the novel rhabdovirus derived from AV_R184 the corresponding full-length amino acid sequences for each gene from all members of *Ledantevirus* were aligned. All alignments were created with mafft version 6.240 using the L-INS-i method (40). Poorly aligned regions were removed using trimAl version 1.2 (41). Phylogenetic trees were inferred in RAxML version 8.2.10 (42) using the LG substitution model (43) with an invariable site plus discrete Gamma model of rate heterogeneity across sites as determined by Modelfinder implemented in IQ-TREE version 1.6.12. Substitution models were selected based on the AIC criterion (44). Support for each node was determined using 1000 bootstrap replicates. Trees were visualised using figtree version 1.4.4 (http://tree.bio.ed.ac.uk/software/figtree/).

### Nested RT-PCR amplification

Plasma RNA was extracted from 140 μl plasma aliquots using QIAamp Viral RNA Mini Kit (Qiagen Cat No 52904). Next, cDNA was generated in a 30 μl reaction containing RNA, reverse primer LEDPOL-51R (5’-CAACGCACATATCCTTCATCATCAGC-3’) and master mix reagents (SuperScript IV (SSIV) Reverse Transcriptase Kit, ThermoFisher 18090050) following manufacturer’s recommendations. Nested PCR amplification was carried out using Platinum™ Taq DNA Polymerase High Fidelity (ThermoFisher cat No 11304029) and the following primer sets aimed to amplify a 5’-half genome sequence of the metagenomic-inferred LDV strain. This fragment contains nucleoprotein, phosphoprotein, matrix, glycoprotein genes, and a small polymerase subgenomic fragment .First round amplification performed with forward primer LEGA5-F (5’-TTTTCTGGTCTTCTCTTTTTCCTACTGAAA-3’) and reverse primer LEDPOL-51R (see sequence above) generates a 5,706 bp fragment. Second round PCR amplification carried out with forward primer LDAN-F (5’-ATGGCTAACGAGACAATTTATCGTTTCTC-3’) and reverse primer LEDPOL-52R (5’-GATGCATTCAACATCAACACCATGTCATG-3’) generates a 5,464 bp fragment. A PCR positive control included an RNA extract from the Le Dantec viral isolate DakHD763 obtained from an infected Senegalese child in 1965. DakHD773, a BSL-2 rhabdovirus was imported as a lyophilized cultured supernatant derived from an experimentally infected mouse, as a kind donation from Dr. Thomas Ksiazek from the University of Texas Medical Branch, Galveston, TX 77555). Experimental positive PCR reactions visualized in agarose gel electrophoresis were sequenced by Sanger chemistry using ABI Big dye® Terminator v3.1 kit and ABI 3500 Genetic Analyser (ThermoFisher) per manufacturer’s recommendations.

### Le Dantec real-time RT-PCR

RNA was extracted from 200µl of plasma using the Agencourt RNAdvance Blood Kit (Beckman Coulter) or the Qiagen viral minikit (see above) according to the manufacturer’s instructions. Extracted RNA was tested directly for Le Dantec virus by real-time PCR or stored at −80°C until use. An in-house developed real-time RT-PCR assay targeting a 145 bp fragment of the LDV glycoprotein gene was develop as a highly sensitive detection assay of the Le Dantec virus in clinical samples. An aliquot of RNA was mixed in a 30 μl reaction volume containing reagents from SuperScript III Platinum One-Step Quantitative RT-PCR System (ThermoFisher Cat No. 11732-020) 0.2 mM forward primer LEDAG-F (5’-GCTTGAAATGCCCTGAAGCT-3’), 0.2 mM reverse primer LEDAG-R (5’-TCACATCTRGTCAACCATCTTGA-3’) and 0.1 mM TaqMan probes labelled with 6-carboxyfluorescein (FAM) LEDAG-FAM (5’-CCATCACCCTATGTTCATCAGGACC-3’) in the presence of 0.05 μM ROX™ Reference Dye. RT-PCR conditions were those recommended by the manufacturer for 50 cycles of amplification in an ABI 7500 Real-Time PCR System (Applied Biosystems).

### LDV-G ELISA

The development of the LDV-G ELISA has been described previously (10). Briefly, the LDV-G transmembrane domain was identified using TMHMM (45). The predicted ectodomain of LDV-G was cloned into the secretory mammalian expression vector pHLSec containing a C-terminal 6xHistidine tag. Human embryonic kidney cells (HEK 293T), maintained in Dulbecco modified Eagle’s medium supplemented with 100 IU/ml penicillin, 100 µg/ml streptomycin, 2 mM glutamine and 10% foetal bovine serum were transfected using Fugene 6 (Promega) and cell supernatant containing secreted LDV glycoprotein was harvested at 48 hours post transfection. Presence of His-tagged protein was confirmed by Western Blotting against 6xHis.

ELISA plates (Thermo Scientific) were coated with rabbit polyclonal anti-6xHis antibody (Abcam) diluted to 1:1000 in basic coating buffer and incubated overnight at 4 °C. The next day plates were blocked with 2.5% BSA in PBS at 37 °C for 1 hour. Cell supernatant containing LDV-G-His or mock protein was applied to the wells followed by human serum diluted 1:50 in blocking buffer or purified IgG for 1 hour at 37 °C. This was followed by goat anti-human IgG peroxidase antibody (Sigma) at room temperature for 1 hour. Wells were washed after each step with 0.1% Tween-20 in PBS. The ELISA reaction was developed using TMB substrate (Thermo Scientific), stopped with 0.16M sulphuric acid and absorbance read at 450 nm using a Pherastar FS plate reader (BMG Labtech). Each sample was tested in a test well containing the truncated LDV-G-His protein as the capture antigen and, to identify nonspecific binding, a mock well with supernatant derived from mock-infected cells transfected with empty plasmid vector. The test result was reported as the OD450 reading from the sample test well divided by that of the sample mock well to give a test/mock OD450 ratio for each sample. For each serum sample the ELISA was repeated on at least three occasions on separate plates. Each plate included known positive and negative control samples, as well as a blocking buffer only sample. All patient samples were heat-inactivated by incubation at 60 °C for 30 minutes before use in the assay.

### Production of VSVΔG*luc* pseudoparticles

For expression of the KEUV, KCV and VAPV glycoproteins (KEUV-G, KCV-G and VAPV-G), complete coding sequences were derived from GenBank accession numbers NC_034540.1, NC_034451.1, and NC_043538.1 and synthesised with the addition of a SalI restriction site and Kozak sequence immediately before the 5’ initiation codon and a NotI restriction site immediately following the 3’ stop codon (GeneWiz/BioBasic). For expression of LDV-G the glycoprotein sequence derived from the Ugandan patient was codon optimised and synthesised (Eurofins genomics). Coding sequences of LDV-G, KEUV-G, VAPV-G and KCV-G were then cloned into the multiple cloning site of mammalian expression vector VR1012 using the SalI and NotI restriction sites, or the NotI and BglII restriction sites in the case of LDV-G, to create VR1012-LDV-G, VR1012-KEUV-G, VR1012-VAPV-G and VR1012-KCV-G. Plasmid pMDG was used to express VSV-G.

For each pseudotype virus 2×10^6^ HEK293T cells were seeded into 10 cm dishes in 10 ml DMEM supplemented with 10% FCS. The next day cells were transfected with the relevant glycoprotein expression plasmid using Fugene 6 (Promega) as per the manufacturer’s instructions. One 10cm dish was not transfected to serve as a no glycoprotein control. Plates were incubated overnight at 37 °C in a 5% CO_2_ atmosphere. The next day plates were removed from the incubator and infected with 15ul of 1.15x 10^7^ TCID50/ml VSVΔG*luc*-VSV-G (corresponding to an approximate MOI of 0.03) and returned to the incubator for 3 hours. Media was discarded and cells washed three times with warmed PBS before the addition of 10 mls DMEM with 10 % FCS. The cells were re-incubated for a further 72 hours after which supernatants containing pseudotype virus were harvested, passed through a 0.45 µm filter (Starlab) and stored at −80 °C before further use.

### Pseudotype-based neutralisation assay

HEK293T cells were grown in a 75 cm^2^ flask until 70% confluent. Media was removed and cells washed with 2 ml trypsin before being resuspended in 8 ml DMEM + 10% FBS. Cells were then plated at a density of 5×10^4^ cells per well in a volume of 50 µl per well in each well of a white 96-well plate.

Serum samples were serially diluted in DMEM supplemented with 10% FBS. Pseudotype preparations were mixed 1:1 with each dilution and incubated at 37 °C for 30 minutes. 50 µl of the pseudoparticle/sera mixture was added to the plated cells in triplicate for a final dilution series ranging from 1:32 to 1:131,072 and a “no sera” control well. Plates were incubated at 37 °C, 5 % CO_2_ for 24 hours before addition of 75 µl Steadylite luciferase substrate solution (Perkin Elmer) to each well. Plates were incubated at room temperature for 10 minutes before luminescence was read using a Chameleon V luminometer (Hidex). Luciferase activity for each dilution was derived from the mean of three plate replicates normalised to the average for the no sera control wells. Percentage neutralisation for each dilution was calculated by subtracting the normalised luciferase activity from 1 and multiplying by 100. The IC50 value for each serum sample and pseudotype combination was determined by interpolation of a four-parameter logistic regression curve between percentage neutralisation and the reciprocal serum dilution. Regression curves were fitted in R version 3.6.3/R Studio version 1.2.5033 using the package drc version 3.0-1 with an upper constraint of 100 using a minimum of 2 biological replicates to fit each curve.

### Immunocytochemistry

The LDV-G sequence was cloned into the expression vector VR1012 with the addition of a C-terminal 6xHistidine tag to generate VR1012-LDV-G-His. Coverslips (Fisher Scientific) were added to each well of a 24-well plate (Corning) and 0.5 ml 70 % IMS added to each well for 20 minutes. IMS was removed and coverslips allowed to air dry before being washed 3 times with 1ml PBS. BHK-21 cells were plated at a density of 1×10^5^ per well and incubated at 37 °C, 5% CO_2_ until 70% confluent. Cells were transfected with VR1012-LDV-G-His using Fugene 6 transfection reagent (Promega) and returned to the incubator for 24-48 hours.

Media was removed and cells fixed with 4% formaldehyde before permeabilization with 200 µl 0.1% Triton-X-100 per well for 5 minutes. Cells were then washed three times with 1000 µl PBS. The primary antibody mixture comprised a rabbit polyclonal to 6X His tag antibody (Abcam) at a dilution of 1:500 mixed with human serum at a dilution of 1:200 with BSA 1% as the diluent (ThermoFisher Scientific). The primary mixture was added to the cells at a volume of 200 µl per well and incubated overnight at 4 °C. The next morning cells were washed three times with PBS and blocked with 1 % BSA for 30 minutes. The secondary antibody mixture was a combination of an Alexa Fluor 488-conjugated goat anti-rabbit IgG secondary antibody (Invitrogen) at a concentration of 4 µg/ml and an Alexa Fluor 594-conjugated goat anti-human IgG secondary antibody (Invitrogen) at a concentration of 5 µg/ml diluted in 1% BSA. The secondary antibody mixture was added to wells at a volume of 200 µl and the plate placed on a shaker at room temperature for 45 minutes before three further 5 minute washes with 1000 µl PBS. Coverslips were stained with Hoechst 33342, at a concentration of 1:2000 in PBS for 15 minutes. Coverslips were then washed three times with 1000 µl PBS and once with ultrapure water before being mounted on glass slides with 2 µl CitiFluor AF1 Mountant solution (Electron Microscopy Sciences). Images were acquired using a LSM 710 confocal microscope (Carl Zeiss Microscopy), and images processed using ZEN Blue software (Carl Zeiss Microscopy).

### Environmental analysis

The R package raster version 3.4-5 was used to extract bioclimatic, geographical and livestock data at the locations of the ANC-2016 study sites. Bioclimatic variables were derived from the Worldclim dataset (46). Tree cover data was extracted from the global forest change dataset (47). Livestock density data was extracted from the UN FAO dataset (https://livestock.geo-wiki.org/home-2/). Elevation was extracted from the STRM 90m DEM Digital Elevation Dataset (https://srtm.csi.cgiar.org/). Variables were averaged across a 10km radius around each study site. Multicollinearity between variables was tested by creating a correlation matrix by the method of Spearman and subsequently a hierarchal clustering dendogram to identify highly correlated groups of variables. A correlation coefficient cut-off of 0.6 was used to group variables. One variable from each group was selected to be included in the global model.

Binomial generalised mixed models were constructed in R version 3.6.3/R Studio version 1.2.5033 using package lme4 version 1.1.26. In addition to bioclimatic variables, patient level age was included in the analysis. To identify the optimal random effects structure an initial model was created combining all fixed effects terms and a random effects term of either site, region or a nested random effect (region/site), with the optimal random effect term selected based on the AICc criterion (48). The best fitting model was then identified using the dredge function package MuMIn version 1.43.17 (49), with the AICc criterion used to select the final model. The response variable was patient level positivity on LDV ELISA, with the final fixed effect explanatory variables being patient age, forest cover, and isothermality (BIO3), with a random effect of site (24 levels). Continuous response variables were normalised. The model was called as glmer(seropositivity) ∼ age + scale(forest_cover,center=(mean forest cover))) + scale(isothermality,center=(mean isothermality)) + (1|site), family=binomial). Model assumptions were tested using the R package DHARMa version 0.4.4 (50). Conditional and marginal R2 measures were computed using R package MuMIn.

## Inclusion statement

The research was conducted as part of a partnership between the MRC-University of Glasgow Centre for Virus Research, the MRC/UVRI & LSHTM Uganda Research Unit and the Uganda Virus Research Institute. The study was designed and conducted by researchers based in Uganda and the UK. Field and laboratory work were performed by trained individuals following a full health and safety risk assessment. Field PPE was employed when handling small mammals (rubber boots, coveralls, goggles and gloves). Laboratory samples were handled in a class 2 biological safety cabinet. The study was reviewed by the UVRI research ethics committee and the UNCST. All sampling and fieldwork protocols relating to wildlife sampling were based on established protocols in use at UVRI. All individuals meeting the criteria for authorship have been included as authors.

## Data Availability

Consensus viral genome and mitochondrial cytochrome B sequences have been submitted to GenBank under accession numbers OQ077988, OR497399 and OR607590. Raw sequencing read files derived from rodent blood have been submitted to NCBI sequence read archive (BioProject ID PRJNA1013481, Biosample accession SAMN37299165). Experimental data relating to LEDV-ELISA and pseudotype-based neutralisation assays will be made available on request to the authors.

---

**Figure S1.**
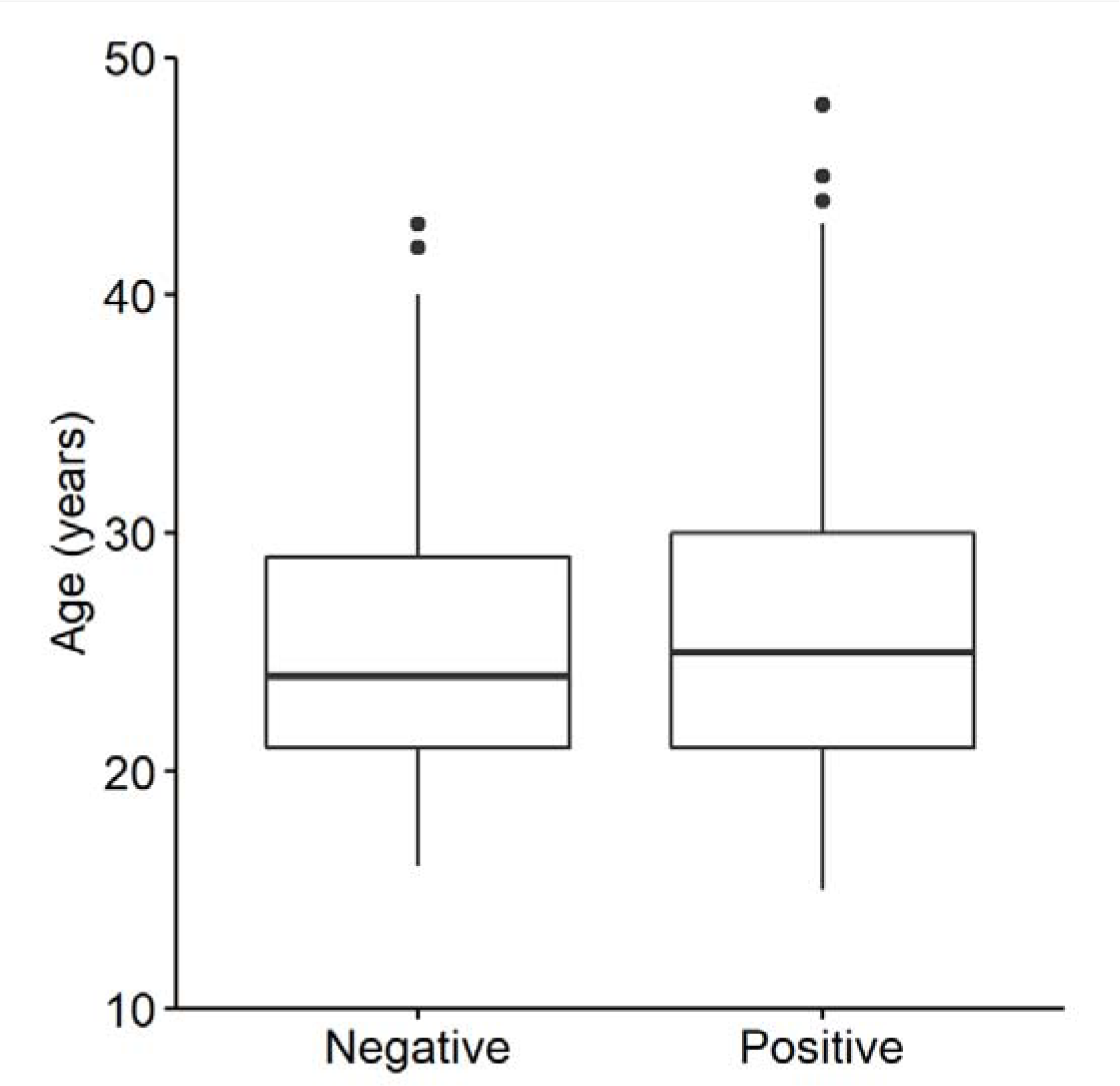
Age by LDV-G ELISA reactivity in the ANC-2016 cohort. Age by seroprevalence as determined by LDV-G ELISA for individuals in the ANC-2016 cohort (n=997). Median age in the seropositive group was 25 years (n=451, IQR=21-30) compared to 24 in the seronegative group (n=546, IQR=21-29), Mann-Whitney U, p=0.016. Centre bars represent the median, box edges the IQR, vertical lines the range (1.5 times the IQR from the upper and lower quartile), and points the outliers.

**Figure S2:**
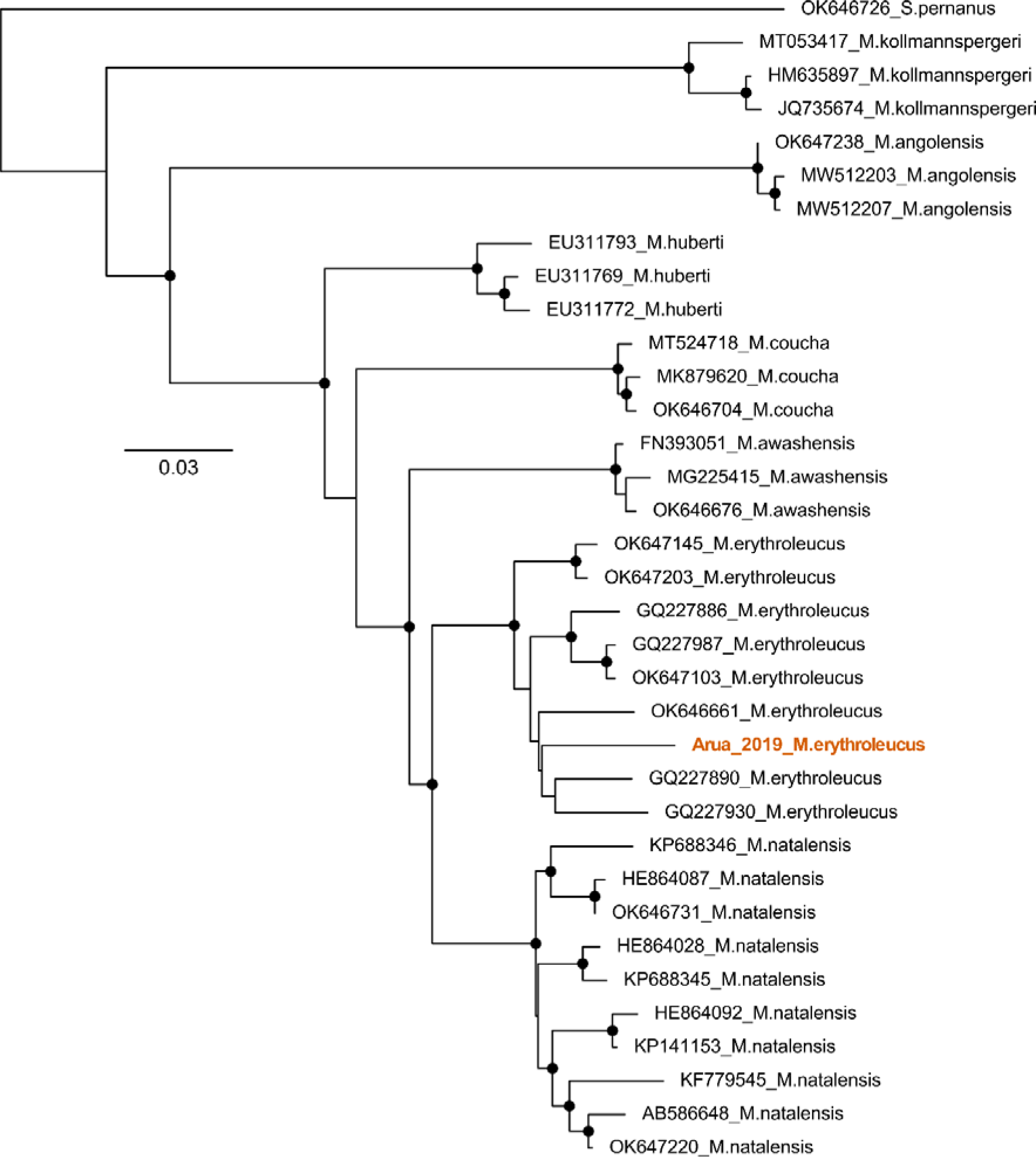
Species identification of *Mastomys erythroleucus* by mitochondrial cytochrome B sequencing. Maximum likelihood phylogeny of mitochondrial cytochrome B sequences derived from representative *Mastomys* species. The sequence derived from the animal in which Odro virus was detected is indicated in orange. *Serengetimys pernanusis* is included as an outgroup. Circles indicate nodes with bootstrap support >70.

**Supplementary Figure S3.**
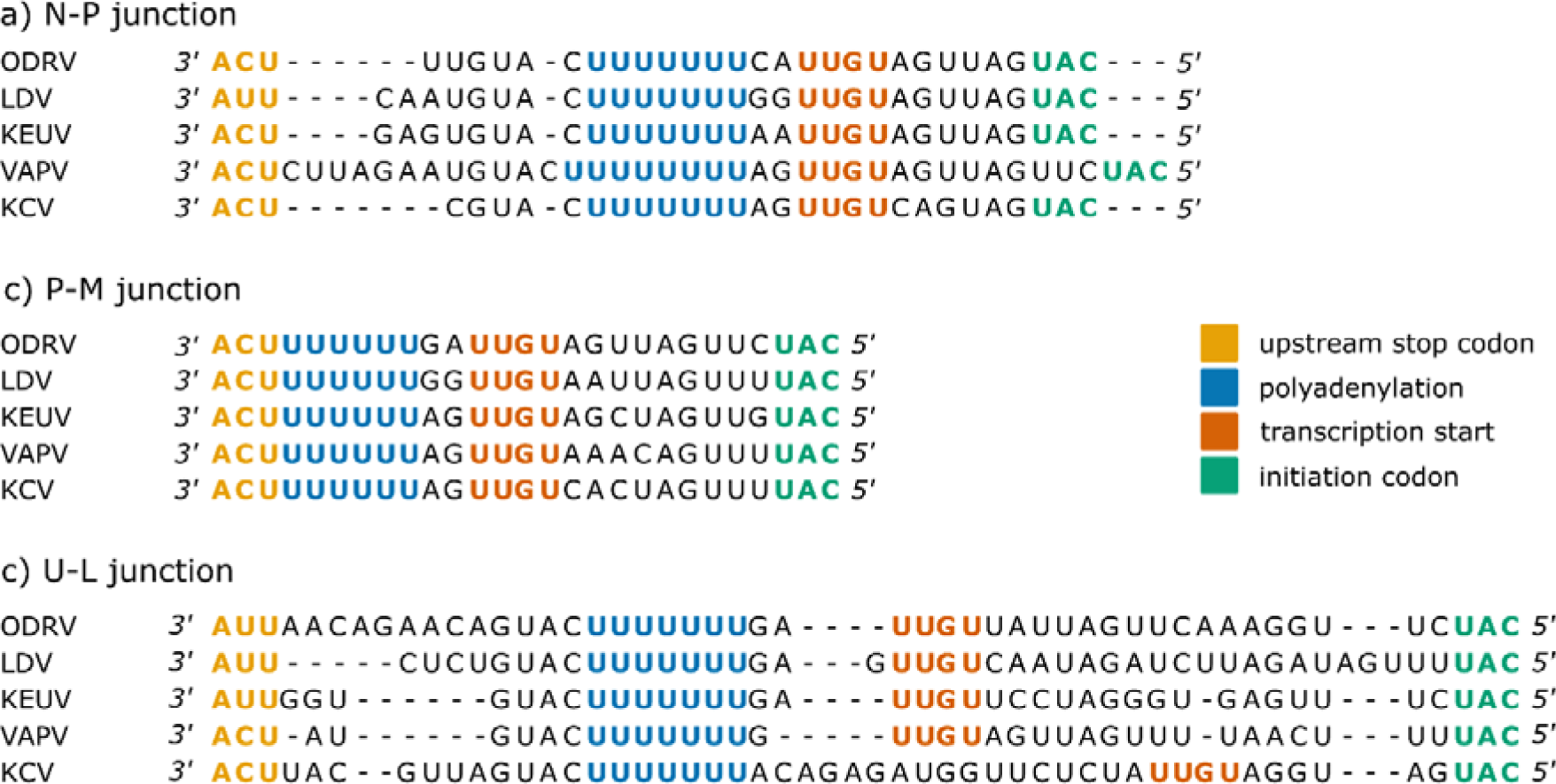
Odro virus shares conserved gene junction regions with other group B ledanteviruses. Genomic gene junction regions of Odro virus compared with other group B ledanteviruses demonstrating the presence of conserved rhabdovirus intergenic polyadenylation and transcription initiation signals. a) nucleoprotein – phosphoprotein junction. b); phosphoprotein matrix protein junction. c) accessory protein – RNA dependant RNA polymerase junction. ODRV; Odro virus, LDV; Le Dantec virus, KEUV, Keuraliba virus, VAPV; Vaprio virus, KCV; Kern Canyon virus.

**Table S1:** Assays employed in the acute febrile illness study (AFI)

| Table S1: Assays employed in the acute febrile illness study (AFI) |  |
| --- | --- |
| Pathogen | Assay |
| Leptospirosis | Leptospirosis IgM lateral flow assay (Lifeassay diagnostics, Cape Town, SA) |
| Brucellosis | Brucella IgM lateral flow assay (Lifeassay diagnostics, Cape Town, SA) |
| Malaria | Thick and thin blood films<br>Rapid 1-2-3 malaria HEMA Express rapid diagnostic kit (Miramar, Florida, USA.) |
| Dengue virus (acute samples) | Dengue NS1 Ag ELISA (Standard Diagnostics Inc. Kyonggido, Korea) |
| Dengue virus (convalescent samples) | Dengue IgM ELISA (Standard Diagnostics Inc. Kyonggido, Korea) |
| Chikungunya | Chikungunya IgM ELISA (Standard Diagnostics Inc. Kyonggido, Korea) |
| O'nyong nyong | In-house IgM ELISA developed at UVRI |
| West Nile Virus | West Nile virus IgM Capture ELISA (FOCUS Diagnostics Cypress, CA) |
| Yellow Fever Virus | In-house ELISA developed at UVRI |
| Typhoid Fever | Acute and convalescent IgM Tubex serology (IDL Biotech, Bromma, Sweden). |
| Rickettsioses | Rickettsia indirect fluorescent (IFA) IgG assay (Focus Diagnostics Cypress, CA) |

**Table S2.** Genomic distance between LDV-Uganda and LDV-Senegal (DakHD763/KM205006)

|  |  |  |  |  |  |  |  |
| --- | --- | --- | --- | --- | --- | --- | --- |
| Region | N | P | M | G | U | L | Intergenic |
| Nucleotide p-distance | 5.25 | 6.84 | 5.68 | 5.75 | 9.23 | 6.38 | 5.17 |
| Amino acid p-distance | 0.24 | 3.86 | 1.87 | 2.62 | 14.06 | 2.35 | - |

**Table S3:** number of rodents by species captured for serum mNGS in Adumi subcounty.

|  |  |
| --- | --- |
| *Aethomys kaiseri* | 17 |
| *Arvicanthus niloticus* | 13 |
| *Crocidura spp.* | 57 |
| *Lemniscomys striatus* | 6 |
| *Mus minutoides* | 7 |
| *Mastomys spp.* | 26 |
| *Rattus rattus* | 64 |
| *Gerbilliscus validus* | 8 |
| *Taterillus emini* | 2 |
| *Thamnomys spp.* | 2 |
| *Zelotomys hildegardeae* | 3 |

**Table S4:** blastx results of *de novo* contigs from sequencing of blood from *Mastomys erythroleucus*.

|  |  |  |  |  |  |
| --- | --- | --- | --- | --- | --- |
| Contig name | Accession of blastx hit | Protein | Virus | Identity (%) | Length (AA) |
| merged_contig3927 | YP_009361873 | polymerase | LDV | 78.2 | 271 |
| merged_contig4193 | YP_009361873 | polymerase | LDV | 87.9 | 471 |
| merged_contig5160 | YP_009361873 | polymerase | LDV | 67.9 | 153 |
| merged_contig5201 | YP_009362198 | glycoprotein | KEUV | 76.5 | 421 |
| merged_contig5497 | YP_009361873 | polymerase | LDV | 86.4 | 140 |
| merged_contig6266 | YP_009361873 | polymerase | LDV | 76.8 | 69 |
| merged_contig7735 | YP_009362195 | nucleoprotein | KEUV | 84.3 | 268 |
| merged_contig8264 | YP_009361873 | polymerase | LDV | 72.1 | 226 |
| merged_contig10327 | YP_009361873 | polymerase | LDV | 84.1 | 531 |
| idba-121_14222 | YP_009362196 | phosphoprotein | KEUV | 63.9 | 72 |
| idba-121_8592 | YP_009362194 | polymerase | LDV | 59.6 | 47 |
| spades_37480 | YP_009361868 | nucleoprotein | LDV | 70.8 | 96 |
| spades_75880 | YP_009361869 | phosphoprotein | LDV | 49.4 | 79 |

**Table S5:** pairwise amino acid distance between Odro virus and other phylogroup B ledanteviruses.

|  |  |  |  |  |
| --- | --- | --- | --- | --- |
| Virus (accession) | Gene |  |  |  |
|  | N | P | G | L |
| KEUV (KM205021) | 21.45 | 50.00 | 24.03 | 21.01 |
| LDV (KM205006) | 21.18 | 46.15 | 24.57 | 18.36 |
| VAPV (MG021441) | 36.46 | 58.19 | 37.71 | 29.37 |
| KCV (KM204992) | 50.40 | 70.39 | 47.79 | 36.46 |

